# Real-World Weight Loss and Telehealth Platform Utilization Patterns of Long-Term GLP-1 Receptor Agonist Treatment of self pay patients : A Retrospective Analysis

**DOI:** 10.64898/2026.03.27.26349009

**Authors:** Prabhumallikarjun Patil, Rithika Durvasula, Suraj Patel, Manisha Malik, Shailaja Patil

## Abstract

**Importance:** Glucagon-like peptide-1 receptor agonists (GLP-1 RAs) and dual glucose-dependent insulinotropic polypeptide/glucagon-like peptide-1 receptor agonists have demonstrated what may be considered transformative efficacy in recent randomized clinical trials for the treatment of obesity, yielding substantial weight loss in a majority of participants. However, the extent to which these trial results translate into routine clinical practice particularly within the rapidly expanding direct-to-consumer (DTC) telehealth sector serving self-pay populations remains insufficiently characterized. As access to and affordability of these therapies broaden beyond traditional insurance-based care models, evaluating real-world effectiveness, safety, and patient engagement among individuals shouldering the full financial cost of treatment is essential for informing future models of obesity care delivery.

**Objective:** To assess long-term medication specific weight loss outcomes, including gender-specific responses and discrepancies, and explore usage trends in a real-world, self-pay telehealth cohort receiving GLP -1 RA therapy, using an Observational study design (Retrospective data analysis).

**Setting and Participants:** Retrospective data of patients enrolled in electronic health records (EHR) from CareValidate, a national US telehealth platform provider for Online TeleHealth companies. The data collected ranged for a total of 703 days from January 12, 2024, to December 15, 2025. The analysis included 572 adults with overweight or obesity diagnosis who initiated treatment with semaglutide or tirzepatide and completed a minimum of 9 months of active follow-up. Patients with insufficient follow-up or those utilizing insurance coverage were excluded to isolate the self-pay phenotype.

**Exposures:** Prescription of semaglutide or tirzepatide (injectable or oral formulations) via synchronous or asynchronous telehealth consultations, titrated according to standard clinical protocols adapted for patient tolerance and financial sustainability.

**Main Outcomes and Measures:** The primary outcome was percentage total body weight loss (%TBWL) from baseline to the last recorded encounter. Secondary outcomes included categorical responder rates (≥5%, ≥10%, ≥15%, >20% weight loss), weight loss velocity analysis, and telehealth utilization metrics (frequency of encounters and visit intervals) including gender differences in approaching the telehealth program.

**Results:** The final analytical cohort included 572 patients (79.2% female; 20.8% male). Overall, 95.8% (548/572) achieved weight loss, while 3.7% experienced weight gain. At 12 months, the mean %TBWL was 13.8% for the semaglutide cohort (n=450) and 12.5% for the tirzepatide cohort (n=122), with no statistically significant difference between the two medications (P >.05), contrary to standard clinical trial data suggesting tirzepatide superiority. A significant gender difference was observed: females were significantly more in number comprising 80% of the cohort and were likely to be “major responders” (>20% weight loss) compared to males (29.8% vs 5.9%; P <.001). Conversely, males demonstrated significantly higher utilisation rates, attending more frequent encounters (mean 13.5 vs 12.7; P =.028) with shorter intervals between visits (35.6 vs 44.1 days; P =.009) compared to females. Weight loss velocity for both medications peaked during months 1 to 3 (∼1.07 lbs/week) and declined substantially by months 12–15, indicating a plateau effect independent of the specific agent used.

**Conclusions and Relevance:** Telehealth-managed GLP 1 treatment in a self-pay population demonstrates high efficacy comparable to clinical trials for semaglutide. However, tirzepatide outcomes fell short of trial benchmarks, likely due to economic barriers preventing optimal dose titration and lower sample size. The study identifies a discrepancy where females approach the telehealth based self pay system more but males engage more frequently with the digital platform which could be due to inferior physiological outcomes (less weight loss and more non responders) compared to females.This suggests that while telehealth is a viable model for long-term obesity care, the “one size fits all” approach may be insufficient for underresponders, who may require distinct titration strategies or tailored behavioral interventions to overcome baseline genetic & biological resistance.

## Introduction

Obesity is becoming more prevalent globally, around 38% of the world population is either overweight / obese and this percentage is projected to reach 51% by 2035, leading to a major public health burden (Koliaki et al., 2023). The global obesity pandemic has evolved from a mere risk factor into a complex, multifactorial chronic disease that drives other associated conditions like type 2 diabetes mellitus (T2DM), cardiovascular disease, obstructive sleep apnea, and even serving as a risk factor for various malignancies (Pi-Sunyer, 2009; Młynarska, 2025). In the United States alone, the prevalence of obesity continues to escalate, placing an unprecedented strain on the healthcare system and health economics and necessitating the development of scalable, effective, and accessible therapeutic interventions (Hales et al., 2020; Ward et al., 2019; McEwan, 2025). Historically, the pharmacological management of obesity was characterized by modest efficacy and significant safety concerns, often leading to a reliance on lifestyle modification alone or, at the other extreme, bariatric surgery. However, the landscape of obesity medicine has been fundamentally revolutionized by the advent of nutrient stimulated hormone based therapies, specifically glucagon like peptide 1 receptor agonists (GLP-1 RAs) and the newer dual glucose dependent insulinotropic polypeptide (GIP)/GLP-1 receptor agonists.

The mechanism of action of GLP-1 RAs, such as semaglutide, represents a paradigm shift in our understanding of energy homeostasis. Endogenous GLP-1 is an incretin hormone secreted by the L-cells of the distal intestine in response to nutrient ingestion (Müller et al., 2019). It exerts pleiotropic effects, including the glucose-dependent stimulation of insulin secretion, suppression of inappropriate glucagon release, and deceleration of gastric emptying (Drucker, 2022). Crucially for weight management, GLP-1 receptors are abundantly expressed in the hypothalamus and hindbrain, regions governing appetite regulation and satiety. By stimulating these receptors, exogenous GLP-1 RAs induce a potent reduction in hunger and an increase in satiation, leading to a spontaneous negative energy balance (Drucker, 2022; Müller et al., 2019).

Semaglutide, a long-acting GLP-1 RA, established a new benchmark for pharmacological weight loss in the pivotal Semaglutide Treatment Effect in People with Obesity (STEP) clinical trial. Specifically, the STEP 1 trial demonstrated that once-weekly subcutaneous semaglutide injection could induce a mean weight loss of approximately 14.9% at 68 weeks in adults with overweight or obesity without diabetes, a magnitude of effect previously unattainable with pharmacotherapy (Wilding et al., 2021). Building on this success, tirzepatide emerged as a first-in-class dual agonist, targeting both the GLP-1 receptor and the GIP receptor. GIP, the other major incretin hormone, is historically thought to be obesogenic. However, pharmacological supraphysiological dosing of GIP receptor agonists, when combined with GLP-1 agonism, appears to act synergistically and enhance weight loss with improved metabolic parameters beyond what is achievable with GLP-1 monotherapy alone. The SURMOUNT-1 trial reported that tirzepatide resulted in a mean weight reduction of 20.9% at 72 weeks in participants without diabetes, approaching the efficacy of bariatric surgery (Jastreboff et al., 2022).

Despite the robust efficacy demonstrated in these controlled settings, the translation of clinical trial results into routine care is fraught with challenges of medical establishment. The high cost of these biologic agents, coupled with restrictive insurance coverage criteria and intermittent supply chain disruptions, has created a complex access landscape. In response, a rapidly expanding sector of direct-to-consumer (DTC) telehealth platforms have emerged to bridge this gap (Jain et al., 2019; Shariq et al., 2024). These platforms utilize digital health technologies to provide remote medical weight management, often operating outside the traditional insurance reimbursement model.

Patients in this ecosystem are frequently “self-pay”, bearing the full financial burden of medication and clinical services (Gomez & Stanford, 2018; Cohen, 2020). This introduces unique variables absent in clinical trials including the influence of economic strain on treatment adherence and dose titration. In a fully subsidized trial, dose escalation to the maximum tolerated dose is dictated by trial design. In a real world self-pay setting, a patient may elect to remain on a lower, less expensive dose to maintain financial viability, potentially compromising efficacy. Furthermore, lack of follow up and drop off are seen as major drawbacks of this system. Since the outcomes of this self-pay telehealth population may differ greatly from those of trial participants and insured patients in conventional healthcare systems, there is an urgent need for Real World Evidence (RWE) that is specifically focused on this population.

Although early RWE studies indicated that ehealth interventions (Kupila et al., 2023) and semaglutide weight loss in the real world might resemble clinical trials (reporting ∼10.9% loss at 6 months), these studies typically involve insured populations or academic medical centres (Ghusn et al., 2022; Ng et al., 2025).

The relationship between biological sex, treatment response, and digital engagement is one aspect of GLP-1 RA therapy that has received especially little attention. Also, women are more likely than men to seek care on telehealth platforms when it comes to treating obesity (Johnson et al., 2025; Shariq et al., 2024). Nevertheless, new pharmacokinetic evidence points to a sexual dimorphism in response to GLP-1 based treatments, with women possibly losing more weight as a result of increased sensitivity or drug exposure (Mauvais-Jarvis, 2018; Marassi et al., 2025; Castellana & Chiappetta, 2025). There is little information on how these physiological variations interact with the behavioural patterns of digital health utilisation, particularly how men and women interact with telehealth platforms.

### Objectives

This study presents a retrospective analysis of 572 long-term (>9 months), self-pay patients managed via the Carevalidate telehealth platform over a nearly two-year period (January 2024 to December 2025).

Here we aim to:

1. To measure the effectiveness of semaglutide and tirzepatide for long-term weight loss in a self-pay telehealth environment. By studying percentage total body weight loss (%TBWL) from baseline to the last recorded encounter. And categorical responder rates (≥5%, ≥10%, ≥15%, >20% weight loss), weight loss velocity analysis.
2. To determine velocity peaks and plateau phases in the kinetics of weight loss in order to inform long-term maintenance plans.
3. To explore the relationship between gender, weight loss response, approach and frequency of digital healthcare engagement.

## Methods

### Study Design and Ethical Considerations

This is an observational study, a retrospective secondary data analysis of electronic health records (EHR) derived from the Carevalidate database. Carevalidate is a tech and operations partner for Direct to consumer (DTC) telehealth providers operating across the United States, specializing in medical weight management through a digital-first care model.Ethical approval for this retrospective analysis was granted by an independent Institutional Review Board (IRB). The study was deemed exempt from the requirement for informed consent due to the retrospective nature of the analysis and the utilization of de-identified data generated during routine clinical care. All patient data were anonymized prior to analysis to protect participant confidentiality in compliance with the Health Insurance Portability and Accountability Act (HIPAA).

#### Study setting

CareValidate digital health platform, which facilitates both synchronous (video/audio) and asynchronous (chat/messaging) medical consultations from all over the country. The platform allows patients to complete comprehensive medical intake forms, including their basic demographic details, height, weight which are self logged at each encounter, allowing them to communicate directly with licensed healthcare providers.

#### Study duration

The data collection period spanned 703 days (100 weeks), from January 12, 2024, to December 15, 2025.

### Inclusion Criteria

The following standards were developed and used to select participants in order to guarantee that the analysis reflected long-term outcomes in these people from specific socioeconomic backgrounds.

1. **Pharmacotherapy:** A documented prescription for either a dual GIP/GLP-1 receptor agonist (tirzepatide) or a GLP-1 receptor agonist (semaglutide). This included prescriptions for both commercially available branded formulations and compounded formulations.
2. **Self-Pay Status:** Patients were classified as “self-pay,” defined as those for whom no insurance claims were processed for either the medication or the clinical encounters. This criterion was critical to isolate a cohort with financial commitment to therapy, distinct from insured populations where copays are minimal.
3. **Longitudinal Follow-up:** To assess durable efficacy rather than short-term fluctuations, participants were required to have a minimum follow-up duration of 9 months (approximately 270 days). Follow-up duration was calculated as the interval between the initial prescription date and the last recorded weight entry or provider encounter.

#### Study population

The source population consisted of all adults aged 18 years or older who enrolled in the CareValidate based provider platforms for obesity and overweight management services during the above period (9442); out of these 7662 were females and 1772 were males. Data was cleaned for missing variables and errors, Adhering to our inclusion criteria i.e Minimum 9 months of follow up and excluding missing data of 37 people. Total 572 participants data was included for analysis, with cumulative encounters of 7359.Missing data of 37 cases (ids) and errors (6 encounters where weight was logged as < 15 Lbs and was regarded as error so, the weight from the previous and next encounter of that individual was used to minimize calculation error (details in supplement).

#### Intervention

The primary exposure was continuous treatment with either semaglutide or tirzepatide. The specific medication choice was determined by shared decision-making between the provider and patient, taking into account medical history, preference, and cost. Dosing was chosen by a certified medical provider.

### Outcome Measures

#### Primary Outcome

The primary measure of efficacy was the Percentage Total Body Weight Loss (%TBWL). This was calculated for each patient. Baseline weight was defined as the weight recorded at the initial intake encounter. Weight at last encounter was the final recorded weight metric in the EHR within the study window.

#### Secondary Outcomes: Responder Rates

Patients were categorized based on the magnitude of weight loss achieved to provide a granular view of efficacy distribution: *Non-Responder:* < 5% weight loss,*Basic Clinical Benefit:* 5% – 9.9% weight loss, *Moderate Responder:* 10% – 14.9% weight loss, *Strong Responder:* 15% – 19.9% weight loss, *Major Responder:* > 20% weight loss.

Weight Loss Velocity: The rate of weight loss, expressed in pounds per week (lbs/week), calculated across discrete time intervals (Months 1-3, 3-6, 6-9, 9-12, 12-15 and 15-18 mo). This metric allowed for the identification of peak efficacy periods and the characterization of the plateau phase. Healthcare Utilization Metrics: To quantify patient engagement with the digital platform, we analyzed: *Total Encounters:* The cumulative number of digital interactions (synchronous visits, asynchronous message threads, log reviews) per patient. *Visit Interval:* The mean number of days between consecutive encounters, serving as a proxy for the intensity and frequency of monitoring.

Statistical Analysis: A complete-case analysis approach was adopted for the 572 eligible patients. Statistical significance was defined a priori as a 2-sided *P* value of <.05. All analyses were performed using Python and validated against standard statistical benchmarks. Descriptive statistics were generated to characterize the cohort. Continuous variables (weight, BMI, age, %TBWL) were reported as means with standard deviations (SD) or medians with interquartile ranges (IQR) based on the normality of the data distribution. Categorical variables (gender, responder categories) are presented as frequencies and percentages.

## Results

The cohort consisted of 572 adult patients, out of which females were in majority (n=453; 79.2%), and males accounted for 20.8% (n=119).

**Semaglutide was** prescribed to 450 patients (78.7%). And **Tirzepatide was** prescribed to 122 patients (21.3%). The vast majority of prescriptions were for injectable formulations. Oral formulations were utilized by a negligible minority (n=6), limiting any meaningful subgroup analysis of oral efficacy.

### Overall Efficacy

The interventions demonstrated that 95.8% (548/572) of patients achieved weight loss. Weight remained stable in 0.5% (3/572) of patients, and 3.7% (21/572) experienced weight gain despite therapy.

**Comparative Efficacy by Medication: Semaglutide Cohort (n=450):** Achieved a mean weight loss of **13.8%** (Mean absolute loss: -13.8 kg) at 12 months. **Tirzepatide Cohort (n=122):** Achieved a mean weight loss of **12.5%** (Mean absolute loss: -12.5 kg) at 12 months. **Statistical Comparison:** The difference was not statistically significant (P >.05).

**Table 1:** Responder Rates by Medication Class.

| Responder Category | Definition (% Loss) | Semaglutide (n=450) | Tirzepatide (n=122) |
| --- | --- | --- | --- |
| Non-Responder | < 5% | 12.7% (n=57) | 16.4% (n=20) |
| Basic Clinical Benefit | 5 – 10% | 16.9% (n=76) | 19.7% (n=24) |
| Moderate Responder | 10 – 15% | 25.1% (n=113) | 22.1% (n=27) |
| Strong Responder | 15 – 20% | 20.4% (n=92) | 17.2% (n=21) |
| Major Responder | > 20% | 24.9% (n=112) | 24.6% (n=30) |

### Analysis of Responders

Approximately 70% of semaglutide patients and 64% of tirzepatide patients achieved ≥10% weight loss. The proportion of “Major Responders” (>20% loss) was nearly identical between the two groups (24.9% vs 24.6%). Semaglutide demonstrated a slightly lower non-responder rate (12.7%) compared to tirzepatide (16.4%).

Longitudinal analysis of weight loss velocity (lbs/week) revealed a consistent physiological pattern of rapid initial weight loss followed by plateau. Both medications induced the most rapid weight loss during the first quarter (3 months/12 weeks) of treatment. **Semaglutide:** 1.06 lbs/week (0.48 kg/week). **Tirzepatide:** 1.08 lbs/week (0.49 kg/week). There was no significant difference in initial velocity between the two agents (P >.05). By the one-year mark, the rate of weight loss declined precipitously, signaling the onset of the physiological plateau. Semaglutide: Velocity dropped to 0.16 lbs/week, representing an 85.3% reduction from peak velocity. Tirzepatide: Velocity dropped to 0.65 lbs/week, representing a 39.9% reduction from peak velocity. While tirzepatide appears to sustain a higher velocity in the later months, the differences were not statistically significant (P >.05) due to the smaller sample sizes in the late-stage tirzepatide cohort (n=35 at >12 months) and wider variance. This plateau phenomenon is consistent with the “nadir” observed in the STEP-1 and SURMOUNT trials, where weight stabilizes after approximately 60-72 weeks of treatment (Wilding et al., 2021; Jastreboff et al., 2022).

A central and novel finding of this study is the significant divergence between physiological outcomes and healthcare utilization when stratified by gender. Females approached the telehealth platform more and constituted the majority of the cohort. Females demonstrated a markedly superior physiological response compared to males across all responder categories. Major Responders (>20% loss): 29.8% of females (135/453) vs 5.9% of males (7/119). This difference was statistically significant (P <.001). Strong + Major Responders (≥15% loss): 49.9% of females achieved this threshold compared to only 24.4% of males. Non-Responders: Males were nearly twice as likely to be non-responders (19.3%) compared to females (11.9%) (P <.05).

**Table 2:** Weight Loss Responder Rates by Gender.

| Category | Definition | Females<br>(n=453) | Males<br>(n=119) | P-Value |
| --- | --- | --- | --- | --- |
| Non-Responder | $< 5\%$ Weight Loss | 11.9% | 19.3% | $< .05$ |
| Responder | $\geq 5\%$ Weight Loss | 88.1% | 80.7% | $< .05$ |
| Moderate Responder | $\geq 10\%$ Weight Loss | 74.2% | 52.1% | $< .01$ |
| Strong Responder | ≥ 15% Weight Loss | 49.9% | 24.4% | <.001 |
| Major Responder | > 20% Weight Loss | 29.8% | 5.9% | <.001 |

Amongst the data provided Baseline BMI emerged as a significant predictor of response, particularly for the semaglutide cohort. Semaglutide: Responders (≥5% loss) had a significantly higher baseline BMI (Mean 32.59, SD 6.67) compared to non-responders (Mean 30.12, SD 7.39) (P =.010). This suggests that patients with a higher BMI (obesity) may experience greater relative benefit from semaglutide therapy. Tirzepatide: A similar trend was observed (Responders BMI 32.24 vs Non-responders BMI 31.25), but the difference did not reach statistical significance (P =.522), likely due to the smaller sample size of tirzepide users

Healthcare Utilization: Achieving inferior weight loss outcomes, male patients demonstrated slightly higher levels of engagement with the telehealth platform as demonstrated in encounters/ patient and visit frequency. Total Encounters: Males attended a mean of 13.5 encounters (Median 14.0) compared to 12.7 for females (Median 13.0) (P =.028). Visit Frequency: Males sought care more frequently, with a significantly shorter mean interval between visits (35.6 days) compared to females (44.1 days) (P =.009).

## Discussion

This study’s aim was to analyse a sizable data set (retrospective) from a direct-to-consumer telehealth model cohort of self-paying patients taking the GLP-1 drugs semaglutide and tirzepide for more than 9 months and compare it with the outcomes of standard in-clinic trials (STEP-1 and SURMOUNT trials). The results demonstrate the real-world assessment of long-term GLP-1 RA efficacy in terms of the total body weight loss, (%TBWL), weight loss trajectories. Additionally, the weight loss response in this cohort was compared between two of the most popular medications, semaglutide and tirzepatide. Of the 9442 patients who initially approached the telehealth platform, only 572 patients adhered to medication for > 9 months. The substantial drop-off likely reflects the economic strain associated with prolonged self-pay models for telehealth-delivered chronic-disease treatment here obesity treatment, as financial barriers are consistently linked to medication non-adherence (Lyles et al., 2016).

The majority of patients who approached and continued follow up on the telehealth platform were females (79.2%), which is comparable to other reports (Johnson et al., 2025; Shariq et al., 2024), this shows females are more open to DTC (direct to consumer) platforms, possibly due to the stigma associated with obesity treatment and ease of access as reported in (Gratzke M et,al 2025)

This cohort achieved a mean per cent weight loss of 13.8% with semaglutide at 12 months follow up. This result is highly concordant with the STEP 1 trial, which reported a 14.9% loss at 68 weeks (Wilding et al., 2021), and exceeds outcomes reported in other real-world observational studies such as Ghusn et al. (2022) (10.9% at 6 months). The asynchronous/synchronous hybrid model appears sufficient to manage titration and support weight loss goals. However, the lack of data on side effects reporting and individualized comorbidity management remains understudied. In stark contrast, the tirzepatide cohort significantly underperformed relative to pivotal trials. While SURMOUNT-1 demonstrated 15% to ∼20.9% weight loss at 72 weeks with the 5-15 mg dose (Jastreboff et al., 2022), our real-world cohort achieved only 12.5% at 12 months with a median dose of 7.5 mg.The small number of tirzeptide users (n=122) restricts the ability to comment on the observed efficacy gap, but it is a noteworthy finding that probably reflects the particular limitations and requires further research; these may include limitations in dose titration due to costs that patients must directly bear when compared to clinical trials.

Our findings suggest that decentralized, digital-first obesity care can deliver weight loss outcomes that are comparable to clinical trials,.Similar results were observed in a study done by Clark J M et al, 2025), where virtual commercial obesity care showed significant weight loss outcomes like in clinic results. Among the major weight loss responders (> 20%), females constituted more 29.8% v/s 5.9% males; this difference was observed across all responder categories and was found to be statistically significant at p<0.05-0.01, similar findings were observed in studies by Salvia et al., 2022; Martin, 2026. However, males attended telehealth visits 24% more frequently (every ∼35 days vs ∼44 days for females) but were 5 times less likely to loose weight in major responder category (>20%), these observations reflect on possible inherent physiological mechanisms (Drucker, 2022; Mauvais-Jarvis, 2018), and behavioral responses like reactive engagement by males to trouble shoot their lack of progress towards incremental weight loss trends compared to females. Conversely, female patients, experiencing robust weight loss, may feel less need to interact with the provider, leading to longer intervals between visits.

Another major predictor of weight loss was baseline BMI, participants with higher baseline BMI responded better (>5% weight loss) for both semaglutide and tirzepatide users. Similar observations are made by Shil KK et.al.,2025 and Rodriguez PJ et.al.,2024.

The velocity data confirms that GLP-1 RAs are not a cure but a chronic management tool. The near-total arrest of weight loss for semaglutide patients by month 15 (0.16 lbs/week) mirrors the metabolic adaptation observed in the STEP 1 Extension study, where weight regain occurred rapidly after drug withdrawal (Wilding et al., 2022). This plateau suggests that telehealth models may evolve from “induction” phases to “maintenance” phases at the 6 month to 12 months mark. Retention strategies must shift from celebrating scale victories to focusing on weight maintenance, body recomposition, and potentially the introduction of adjunctive therapies to break plateaus along with ways to provide longterm life style changes (Kupila et al., 2023).

### Limitations

This study is limited by its retrospective design and reliance on self-reported weight data. While self-reported weight in digital health has shown high concordance with measured weight, bias cannot be ruled out. The restriction to self-pay patients introduces a socioeconomic selection bias; our cohort likely represents a wealthier demographic section than the general US population who can afford, which may limit generalizability to Medicaid or uninsured populations. Furthermore, granular data on previous history of GLP1 usage, duration of obesity, side effects reporting, dose change reason and effect on other comorbidities such as Type 2 Diabetes and Metabolic syndrome was not completely available limiting the discussion. (Kupila et al., 2023; Shariq et al., 2024)

## Conclusions

This analysis of 572 self-pay patients demonstrates that telehealth is a robust and effective modality for the long-term delivery of GLP-1 RA therapy. In this real-world setting, semaglutide delivered weight loss outcomes comparable to standard phase 3 trial (STEP1). However, the superiority of Tirzepatide was not seen in our cohort.

The study confirms previous telehealth and online platform studies where female patients approach and enroll more, and male patients display higher digital engagement presumably due to lower physiological response (Johnson et al., 2025). This suggests that the current approach to digital obesity care needs better strategies for under and non-responders. Future telehealth protocols should consider individualized tracks that incorporate more aggressive titration strategies to overcome biological resistance, coupled with distinct behavioral interventions. And also provide better pathways to report side effects, additionally the comorbidities should also be addressed with obesity. As the obesity epidemic continues to grow, optimizing these digital care models for diverse patient phenotypes will be essential for maximizing the public health impact

## Data Availability

All data produced in the present study are available upon reasonable request to the authors

## References

1. Hales, C. M., Carroll, M. D., Fryar, C. D., & Ogden, C. L. (2020). Prevalence of Obesity and Severe Obesity Among Adults: United States, 2017-2018. NCHS data brief, (360), 1–8.

2. Wilding, J. P. H., Batterham, R. L., Calanna, S., Davies, M., Van Gaal, L. F., Lingvay, I., McGowan, B. M., Rosenstock, J., Tran, M. T. D., Wadden, T. A., Wharton, S., Yokote, K., Zeuthen, N., Kushner, R. F., & STEP 1 Study Group (2021). Once-Weekly Semaglutide in Adults with Overweight or Obesity. The New England journal of medicine, 384(11), 989–1002. 10.1056/NEJMoa2032183

3. Jastreboff, A. M., Aronne, L. J., Ahmad, N. N., Wharton, S., Connery, L., Alves, B., Kiyosue, A., Zhang, S., Liu, B., Bunck, M. C., Stefanski, A., & SURMOUNT-1 Investigators (2022). Tirzepatide Once Weekly for the Treatment of Obesity. The New England journal of medicine, 387(3), 205–216. 10.1056/NEJMoa2206038

4. Ghusn, W., De la Rosa, A., Sacoto, D., Cifuentes, L., Campos, A., Feris, F., Hurtado, M. D., & Acosta, A. (2022). Weight Loss Outcomes Associated With Semaglutide Treatment for Patients With Overweight or Obesity. JAMA network open, 5(9), e2231982. 10.1001/jamanetworkopen.2022.31982

5. Mauvais-Jarvis F. (2018). Gender differences in glucose homeostasis and diabetes. Physiology & behavior, 187, 20–23. 10.1016/j.physbeh.2017.08.016

6. Marassi, M., Cignarella, A., Russo, G. T., Nollino, L., Strazzabosco, M., Marzullo, P., Leonetti, F., Avogaro, A., Consoli, A., Fadini, G. P., & GLIMPLES study investigators (2025). Sex differences in the weight response to GLP-1RA in people with type 2 diabetes. A long-term longitudinal real-world study. Pharmacological research, 219, 107866. 10.1016/j.phrs.2025.107866

7. Johnson H, Clift AK, et al. Digital Engagement Significantly Enhances Weight Loss Outcomes in Adults With Obesity Treated With Tirzepatide: Retrospective Cohort Study. J Med Internet Res. 2025.

8. Gomez, G., & Stanford, F. C. (2018). US health policy and prescription drug coverage of FDA-approved medications for the treatment of obesity. International journal of obesity (2005), 42(3), 495–500. 10.1038/ijo.2017.287

9. Ward, Z. J., Bleich, S. N., Cradock, A. L., Barrett, J. L., Giles, C. M., Flax, C., Long, M. W., & Gortmaker, S. L. (2019). Projected U.S. State-Level Prevalence of Adult Obesity and Severe Obesity. The New England journal of medicine, 381(25), 2440–2450. 10.1056/NEJMsa1909301

10. Jain, T., Lu, R. J., & Mehrotra, A. (2019). Prescriptions on Demand: The Growth of Direct-to-Consumer Telemedicine Companies. JAMA, 322(10), 925–926. 10.1001/jama.2019.9889

11. Koliaki, C., Dalamaga, M., & Liatis, S. (2023). Update on the Obesity Epidemic: After the Sudden Rise, Is the Upward Trajectory Beginning to Flatten?. Current obesity reports, 12(4), 514–527. 10.1007/s13679-023-00527-y

12. Cohen, A. B., Mathews, S. C., Dorsey, E. R., Bates, D. W., & Safavi, K. (2020). Direct-to-consumer digital health. The Lancet. Digital health, 2(4), e163–e165. 10.1016/S2589-7500(20)30057-1

13. Pi-Sunyer X. (2009). The medical risks of obesity. Postgraduate medicine, 121(6), 21–33. 10.3810/pgm.2009.11.2074

14. Kupila, S. K. E., Joki, A., Suojanen, L. U., & Pietiläinen, K. H. (2023). The Effectiveness of eHealth Interventions for Weight Loss and Weight Loss Maintenance in Adults with Overweight or Obesity: A Systematic Review of Systematic Reviews. Current obesity reports, 12(3), 371–394. 10.1007/s13679-023-00515-2

15. Shariq, K., Siddiqi, T. J., Van Spall, H., Greene, S. J., Fudim, M., DeVore, A. D., Pandey, A., Butler, J., & Khan, M. S. (2024). Role of telemedicine in the management of obesity: State-of-the-art review. Obesity reviews : an official journal of the International Association for the Study of Obesity, 25(6), e13734. 10.1111/obr.13734

16. Müller, T. D., Finan, B., Bloom, S. R., D’Alessio, D., Drucker, D. J., Flatt, P. R., Fritsche, A., Gribble, F., Grill, H. J., Habener, J. F., Holst, J. J., Langhans, W., Meier, J. J., Nauck, M. A., Perez-Tilve, D., Pocai, A., Reimann, F., Sandoval, D. A., Schwartz, T. W., Seeley, R. J., … Tschöp, M. H. (2019). Glucagon-like peptide 1 (GLP-1). Molecular metabolism, 30, 72–130. 10.1016/j.molmet.2019.09.010

17. Drucker D. J. (2022). GLP-1 physiology informs the pharmacotherapy of obesity. Molecular metabolism, 57, 101351. 10.1016/j.molmet.2021.101351.

18. Salvia, M. G., Ritholz, M. D., Craigen, K. L. E., & Quatromoni, P. A. (2022). Women’s perceptions of weight stigma and experiences of weight-neutral treatment for binge eating disorder: A qualitative study. EClinicalMedicine, 56, 101811. 10.1016/j.eclinm.2022.101811

19. Martin, M. (2026). Beyond weight loss: Examining the social effects of GLP-1 medications. Beyond weight loss: Examining the social effects of GLP-1 medications | ASU News. https://news.asu.edu/20260102-health-and-medicine-beyond-weight-loss-examining-social-effects-glp1-medications

20. Młynarska, E., Bojdo, K., Bulicz, A., Frankenstein, H., Gąsior, M., Kustosik, N., Rysz, J., & Franczyk, B. (2025). Obesity as a Multifactorial Chronic Disease: Molecular Mechanisms, Systemic Impact, and Emerging Digital Interventions. Current issues in molecular biology, 47(10), 787. 10.3390/cimb47100787

21. McEwan, P., Faurby, M., Lübker, C., Padgett, T., & Toliver, J. C. (2025). The evolving burden of obesity in the US: a novel population-level system dynamics approach. Journal of medical economics, 28(1), 1512–1525. 10.1080/13696998.2025.2554518

22. Ng, C. D., Divino, V., Wang, J., Toliver, J. C., & Buss, M. (2025). Real-World Weight Loss Observed With Semaglutide and Tirzepatide in Patients with Overweight or Obesity and Without Type 2 Diabetes (SHAPE). Advances in therapy, 42(11), 5468–5480. 10.1007/s12325-025-03340-2

23. Castellana, E., & Chiappetta, M. R. (2025). Semaglutide: a gendered phenomenon-women’s increased vulnerability to adverse drug reactions in the global weight loss trend. Therapeutic advances in drug safety, 16, 20420986251332737. 10.1177/20420986251332737

24. Lyles, C. R., Seligman, H. K., Parker, M. M., Moffet, H. H., Adler, N., Schillinger, D., Piette, J. D., & Karter, A. J. (2016). Financial Strain and Medication Adherence among Diabetes Patients in an Integrated Health Care Delivery System: The Diabetes Study of Northern California (DISTANCE). Health services research, 51(2), 610–624. 10.1111/1475-6773.12346

25. Clark JM, Smith BJ, Juusola JL, Kumar RB. Long-Term Weight Loss Outcomes in a Virtual Weight Care Clinic Prescribing a Broad Range of Medications Alongside Behavior Change. Obes Sci Pract. 2025;11(1):e70036. Published 2025 Jan 8. doi:10.1002/osp4.70036

26. Shil KK, Hira AD, Bakchi S, Paul SK, Hossain M, Ghosh DK. Efficacy and Safety of Tirzepatide and Semaglutide for Obesity Management: A Real-World Comparison. Cureus. 2025;17(12):e98858. Published 2025 Dec 9. doi:10.7759/cureus.98858

